# The effect of 18 months lifestyle intervention on brain age assessed with resting-state functional connectivity

**DOI:** 10.1101/2022.09.21.22280182

**Authors:** Gidon Levakov, Alon Kaplan, Anat Yaskolka Meir, Ehud Rinott, Gal Tsaban, Hila Zelicha, Matthias Blüher, Uta Ceglarek, Michael Stumvoll, Ilan Shelef, Galia Avidan, Iris Shai

**Author notes:** Corresponding author: Gidon Levakov, Department of Brain and Cognitive Sciences, Ben-Gurion University of the Negev, Beer-Sheva, Israel. Authorship note: G. Levakov and A. Kaplan share first authorship and G. Avidan & I. Shai share senior authorship.

## Abstract

Obesity negatively impacts multiple bodily systems, including the central nervous system. Retrospective studies that estimated chronological age from neuroimaging have found accelerated brain aging in obesity, but it is unclear how this estimation would be affected by lifestyle intervention. In a sub-study of 102 participants of the DIRECT-PLUS (dietary-intervention-randomized-controlled-trial polyphenol-unprocessed) trial, we tested the effect of 18 months of lifestyle intervention on predicted brain age, based on MRI-assessed resting-state functional connectivity (RSFC). We further examined how dynamics in multiple health factors, including anthropometric measurements, blood biomarkers, and fat deposition, can account for changes in brain age. To establish our method, we first demonstrated that our model could successfully predict chronological age from RSFC in three cohorts (n=291;358;102). We then found that among the DIRECT-PLUS participants, 1% of body weight loss resulted in an 8.9 months attenuation of brain age. Attenuation of brain age was significantly associated with improved liver biomarkers, decreased liver fat, and visceral and deep subcutaneous adipose tissues after 18m of intervention. Finally, we showed that lower consumption of processed food, sweets, and beverages were associated with attenuated brain age. These results suggest that lifestyle intervention has beneficial effects on the trajectory of brain aging.

## 1. Introduction

Brain aging is a complex, multifaceted process with various manifestations in different periods of the human lifespan, brain regions, and imaging modalities^1,2^. Nevertheless, reducing this complex process to a single scalar, the predicted brain age, may capture multiple conditions and risk factors associated with deviation from the normal aging trajectory^3^. Brain age estimation is typically done by predicting chronological age from neuroimaging data in a healthy training group of subjects and applying the fitted model to a new, unseen individual. This procedure enables estimating a measure of brain age independent of the individual’s chronological age. Over-estimation of brain age, in relation to chronological age, is observed in several neurological conditions such as mild cognitive impairment, Alzheimer’s disease (AD), schizophrenia, and depression^4–6^, and is associated with an increase in mortality rate^7^. Similarly, over-estimation of brain age was also found in obesity^8–10^, suggesting that the brain age framework may provide a powerful tool for assessing accelerated brain aging due to excessive weight. Critically, it is unclear whether dietary lifestyle interventions may have a beneficial, attenuative effect on the brain aging process.

Obesity is associated with multiple adverse health impacts also observed in normal aging^11,12^. These comorbidities of obesity and typical aging include the risk of cardiovascular disease^13^, inflammation^14^, type 2 diabetes^15^, DNA damage^16,17^, and neurodegenerative processes^18^. The link between excessive weight and neuronal damage is likely mediated by adiposity, metabolic dysfunction, and alteration in the gut microbiome^19,20^. These, in turn, promote inflammatory metabolic processes in the central nervous system^21^. Accordingly, reduction in gray and white matter volume^22,23^, changes in brain connectivity^24,25^, cognitive impairment^26^, and the prevalence of dementia^27^ were all associated with midlife obesity. These anatomical^2^, functional^28^, and behavioral^29^ findings are also observed during normal aging. An increase in life expectancy^30^ along with a sharp growth in obesity rates^31^ elicit the need to characterize, treat and perhaps prevent obesity-related brain aging.

We previously found that weight loss, glycemic control and lowering of blood pressure, as well as increment in polyphenols rich food, were associated with an attenuation in brain atrophy ^32^. Here, as a sub-study of the Dietary Intervention Randomized Controlled Trial Polyphenols Unprocessed Study (DIRECT PLUS^33^), we examined the effect of 18 months of lifestyle intervention on brain aging attenuation (Fig. 1). We assessed brain age based on resting-state functional connectivity (RSFC) taken before and after the intervention. Brain aging attenuation was quantified as the difference between the expected and observed brain age after the intervention. We trained and validated the age prediction model using two separate cohorts (n=291^34^, 358^35,36^), then applied it to our group of participants from the DIRECT-PLUS (n=102). We hypothesized that a successful reduction in anthropometric measurements following the intervention would attenuate brain aging. We then examined how multiple clinical outcomes, including liver, glycemic, lipids, and MRI fat deposition markers, would be related to attenuated brain aging. Finally, we report the correlation between brain age attenuation and changes in reported food consumption. To the best of our knowledge, this is one of the first studies that examined the beneficial effect of lifestyle interventions on the brain aging trajectory in humans, assessed by resting-state fMRI.

**Fig. 1.**
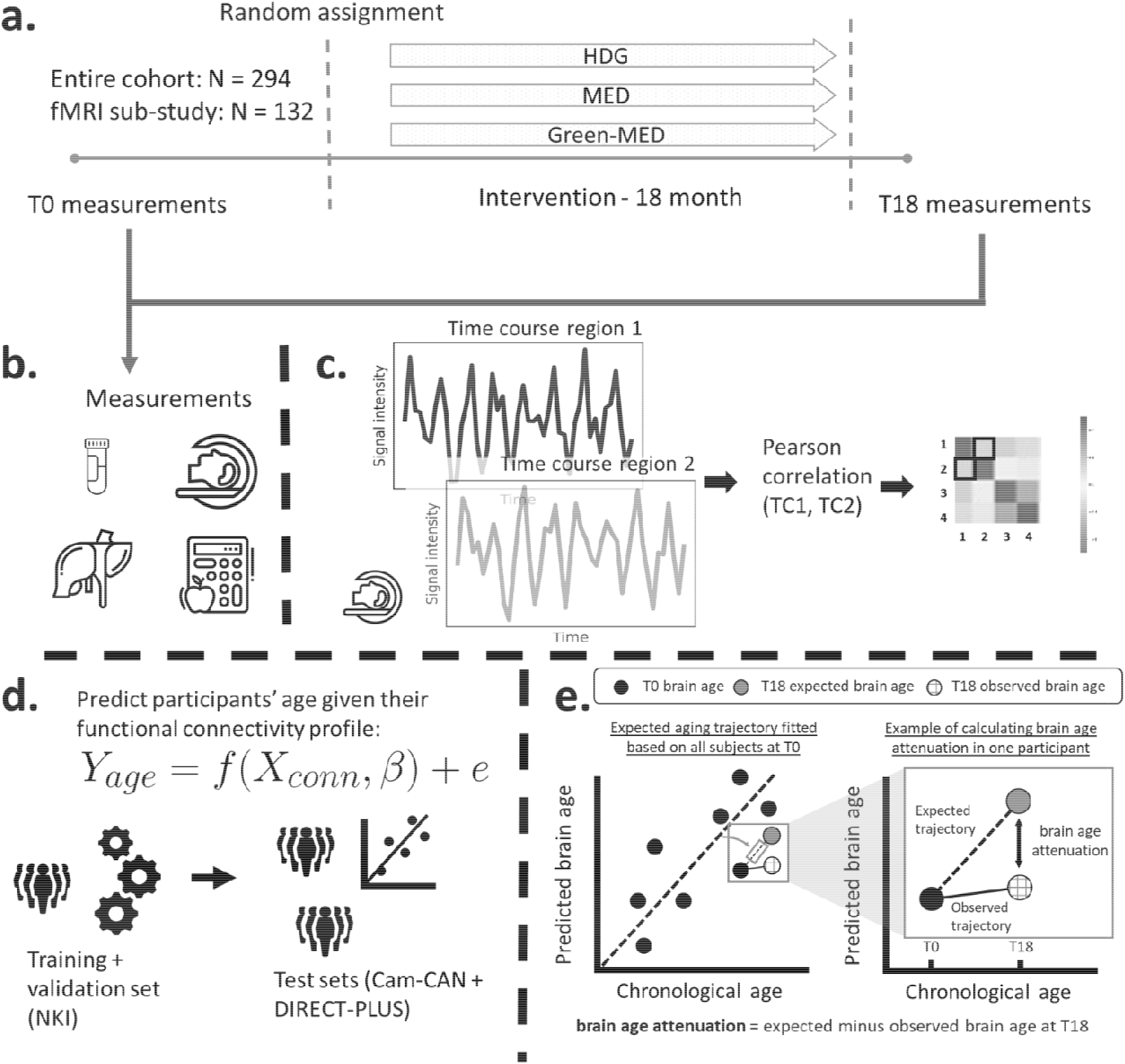
Study design and workflow. The DIRECT-PLUS trial examined the effect of an 18-month lifestyle intervention on adiposity, cardiometabolic, and brain health across intervention groups. (a) Participants in the functional connectivity sub-study (N=132) completed the baseline measurements at T0. They were randomly assigned to three intervention groups: healthy dietary guidelines (HDG), an active control group, Mediterranean diet (MED), and green-MED. All groups were combined with physical activity (PA). Eighteen months following intervention onset, all measurements were retaken (T18). (b) Measurements included anthropometric measurements, blood biomarkers, fat deposition, and structural and functional brain imaging. (c) Functional brain imaging was conducted while subjects were at rest and was used to estimate resting state functional connectivity (RSFC). RSFC measures the correlation between the time series of pairs of brain regions. (d) We fitted a linear support vector regression to predict chronological age from all pairwise correlations. We fitted the model on the NKI data set, then tested and applied it to the Cam-CAN and the DIRECT-PLUS data. (e, left scatter plot) Based on the T0 data, we first computed the expected aging trajectory as the linear relation between the chronological and predicted age of all subjects. The fitted line represents the increase in the predicted age in relation to chronological age in the absence of an intervention. (e, right scatter plot) The fitted line was used to estimate the expected brain age at T18, given each participant’s T0 brain age and the time passed between the T0 and T18 MRI scans. We computed the observed brain age by applying the brain age model to the T18 scans. Brain age attenuation was calculated as the expected brain age minus the observed at T18.

## 2. Results

### 2.1 Brain age estimation

To estimate chronological age from RSFC, we utilized data from 649 participants from two separate cohorts for the brain age model training, validation, and testing. We predicted chronological age from functional connectivity among the 100 nodes of the Schaefer brain atlas^3^ (4950 edges) using a linear support vector regression model. The model was first trained and validated on 291 participants from the Nathan Kline Institute dataset (NKI^34^; n=291) using 5-fold cross-validation. As expected, a positive correlation was found between the predicted and observed age (r=0.439, p<0.001; MAE=8.544, p<0.001). Next, we retrained the model on the entire sample and tested it in an independent sample from the Cambridge Centre for Ageing Neuroscience dataset (Cam-CAN^35^; n=358) again, yielding a positive correlation between the predicted and observed age (r=0.290, p<0.001; MAE=11.402, p=0.005). Finally, we used the fitted model to estimate the brain age within the DIRECT-PLUS cohort. Of the 132 subjects that participated in the fMRI sub-study, 102 were included in all analyses after exclusions due to excessive in-scanner motion (23% omitted; Methods 4.5). The predicted brain age and observed chronological age were correlated (r=0.244, p=0.013; MAE=8.337, p<0.001; Fig. 2), reproducing the results found within the two other datasets. An analysis of the contribution of individual nodes to brain age prediction is provided in the supplementary information (SI) section 10.1.

**Fig. 2.**
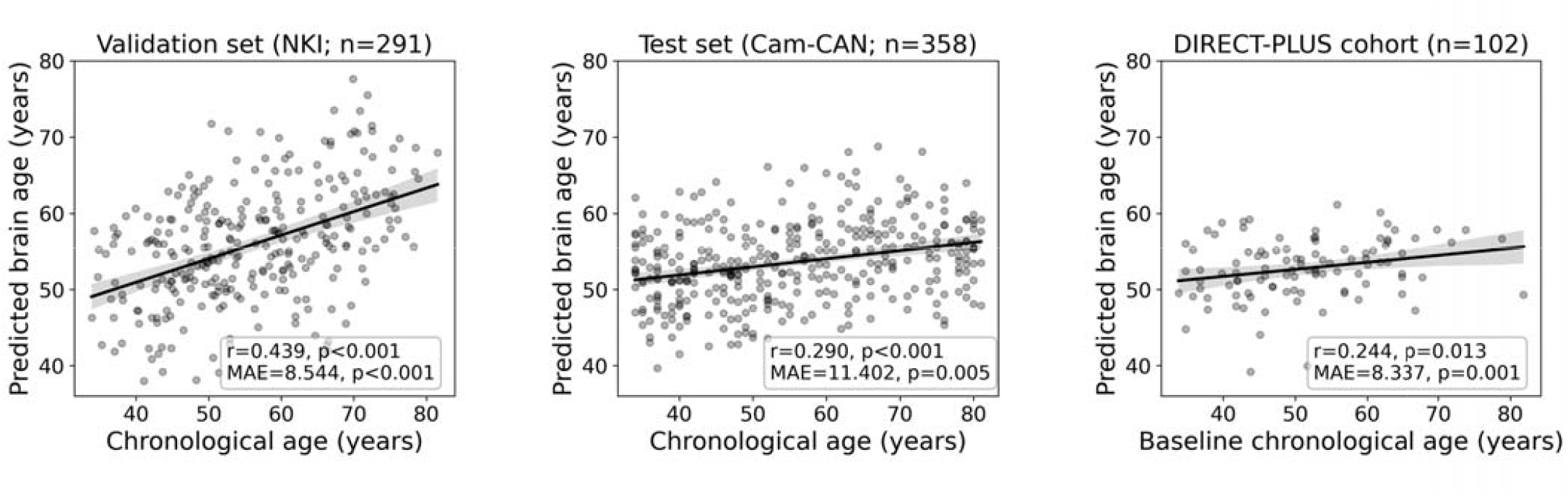
Prediction accuracy within the validation and test cohorts. The scatter plots depict the data points and regression line between the predicted (y-axis) and observed (x-axis) age. The predicted-observed correlation is presented for the validation data (left), the Can-CAN test data (middle), and the DIRECT-PLUS data at baseline. The shaded area around the regression lines represents a 95% confidence interval estimated using bootstrapping. Pearson’s correlation, MAE (mean absolute error), and corresponding p values are shown at the bottom of each plot.

### 2.2 Baseline characteristics

Baseline characteristics among the 102 participants with valid RSFC MRI scans are presented in **Table 1** (see SI Table 1 for additional measures). The mean participant age was 51.5±10.5 years (median=50.6, range 33.9-81.9), and 91.2% were men. The mean body mass index (BMI) and waist circumference (WC) were 30.1±2.5 kg/m^2^ and 107.1±6.6 cm, respectively. The mean baseline predicted brain age by RSFC was 52.8±4 years. At baseline, predicted brain age was associated with chronological age (r=0.24, p=0.013), chemerin (r=0.25, p=0.012), fibroblast growth factor 21 [(FGF21), r=0.21, p=0.034] and with obesity-associated measurements obtained by MRI including visceral abdominal tissue (VAT): r=0.33, p<0.001 and superficial subcutaneous fat (SSC): r=-0.28, p=0.005). Predicted brain age at baseline was also associated with decreased hippocampal occupancy score (HOC), an anatomical measurement of brain atrophy (r=-0.255, p=0.010).

**Table 1.**
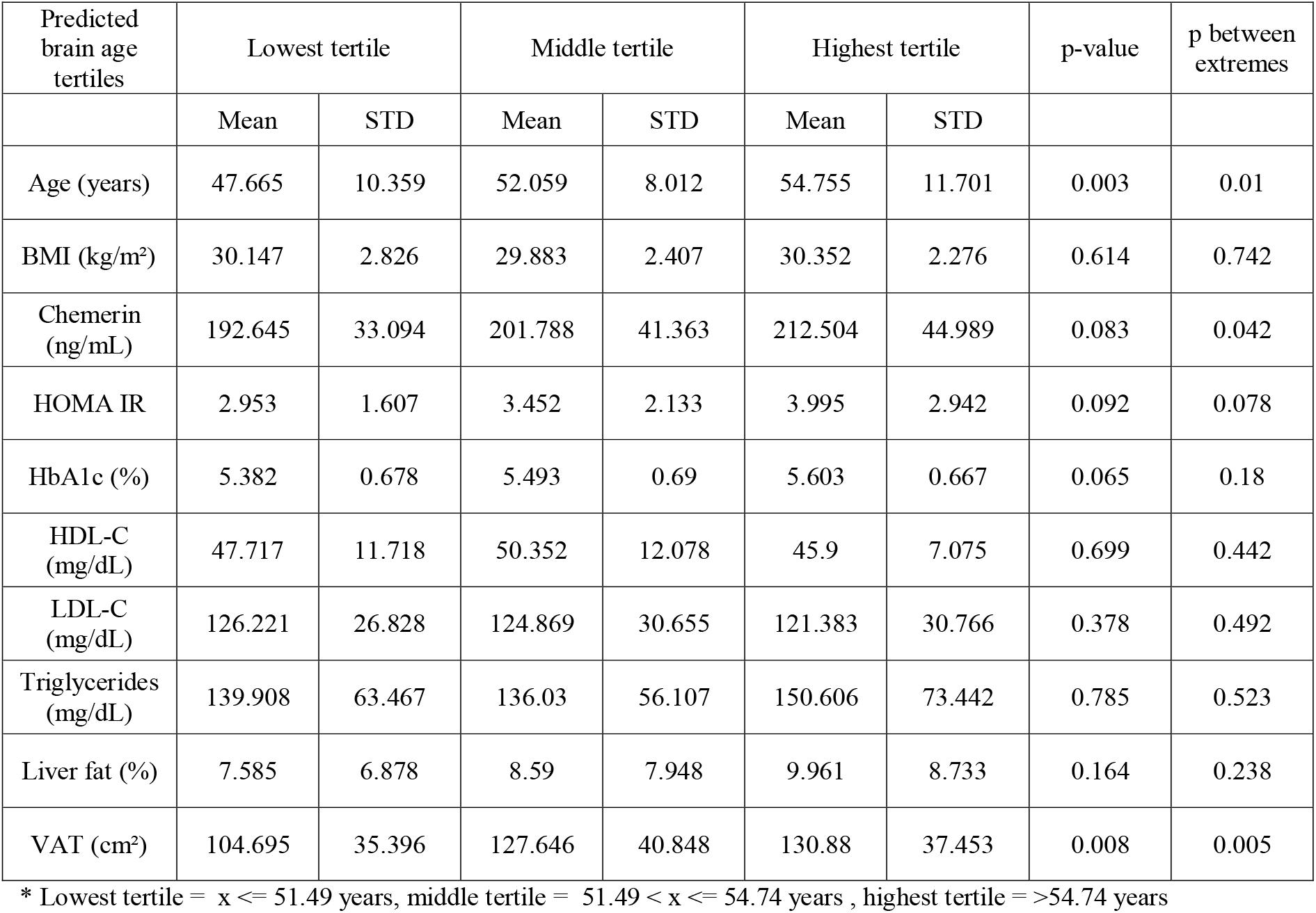
Baseline characteristics according to the baseline predicted brain age tertiles.

### 2.3 The relation between successful lifestyle intervention and attenuation of functional brain aging

Our primary hypothesis was that success in lifestyle intervention, as assessed by anthropometric measurements, will attenuate functional brain aging. Brain aging attenuation was quantified as the difference between the expected and observed brain age at T18 (Fig. 1e). Following 18 months of lifestyle intervention, participants showed a reduction of 0.76 (±1.86) units in BMI on average, 2.31 (±5.61) kg reduction in weight, and 5.39 (±5.89) cm reduction in waist circumference. These constitute a -6.45% ±(5.60%) and -4.35% ±(5.86%) percent reduction from baseline for waist circumference and BMI and weight, respectively. Additionally, at T18, the observed age was lower than expected in 56.8% of the subjects, while the opposite was found in 43.1% of the subjects (X^2^=1.922, p=0.166; see Fig. 3, top). Importantly, we found a correlation between ΔBMI and brain age attenuation such that participants that showed a decrease in BMI also exhibited attenuated brain aging (r=.319, p<0.001; Fig. 3, bottom). Specifically, one percent of BMI or weight loss resulted in an .9 months attenuation of brain age. Similar results were found with Δ bodyweight (r=.319, p<0.001) and Δ waist circumference (r=.198, p=0.046; Fig. 4). The correlations to ΔBMI and Δweight were significant after correcting for age and baseline brain age (p<0.05 for all), while the correlation to waist circumference showed only a trend (r>.171, ps <0.079 for all).

**Fig. 3.**
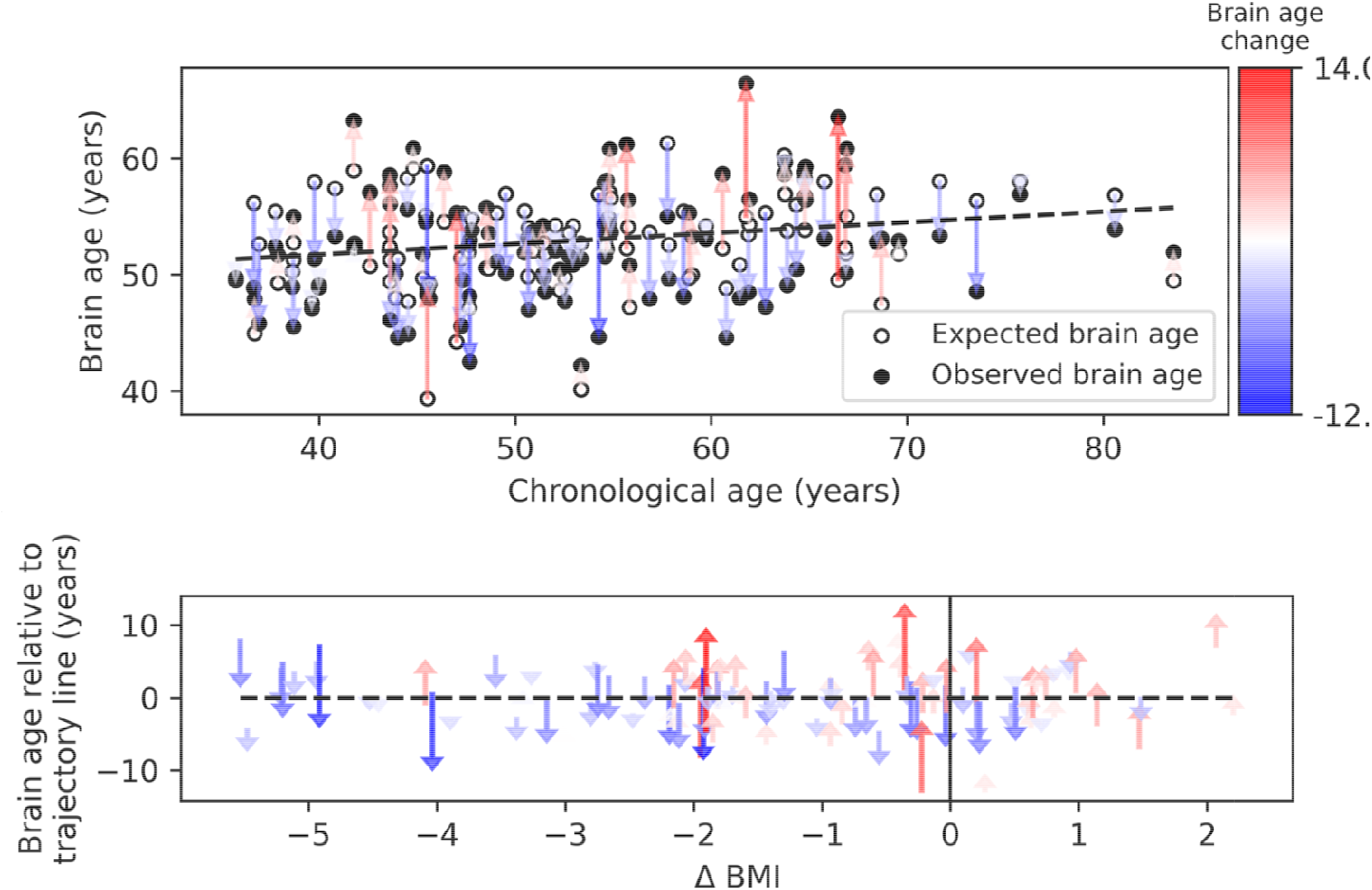
Observed compared to expected brain age at T18. The upper panel depicts the chronological age (x-axis) and the observed (empty circles) and expected (full circles) brain age (y-axis) of each subject. The dashed line represents the expected brain age trajectory fitted based on the T0 data (see regression line in Fig. 1e, left). Arrows point from the expected to the observed age of a single individual, corresponding to brain age attenuation. Arrows’ colors correspond to the extent of brain age attenuation (blue shades indicate attenuation, red shades indicate an acceleration in brain age). The observed age was lower than expected in 56.8% of the subjects, while the opposite was found in 43.1% (X2=1.922, p=0.166). In the lower panel, arrows were reordered by subjects’ BMI change over the 18 months of intervention. A significant correlation was found between the BMI and brain age change (r=.319, p<0.001). This is evident in the graph, such that most of the blue arrows are located on the left side of the x-axis (negative values), and most of the red arrows appear on the right side (positive values).

**Fig. 4.**
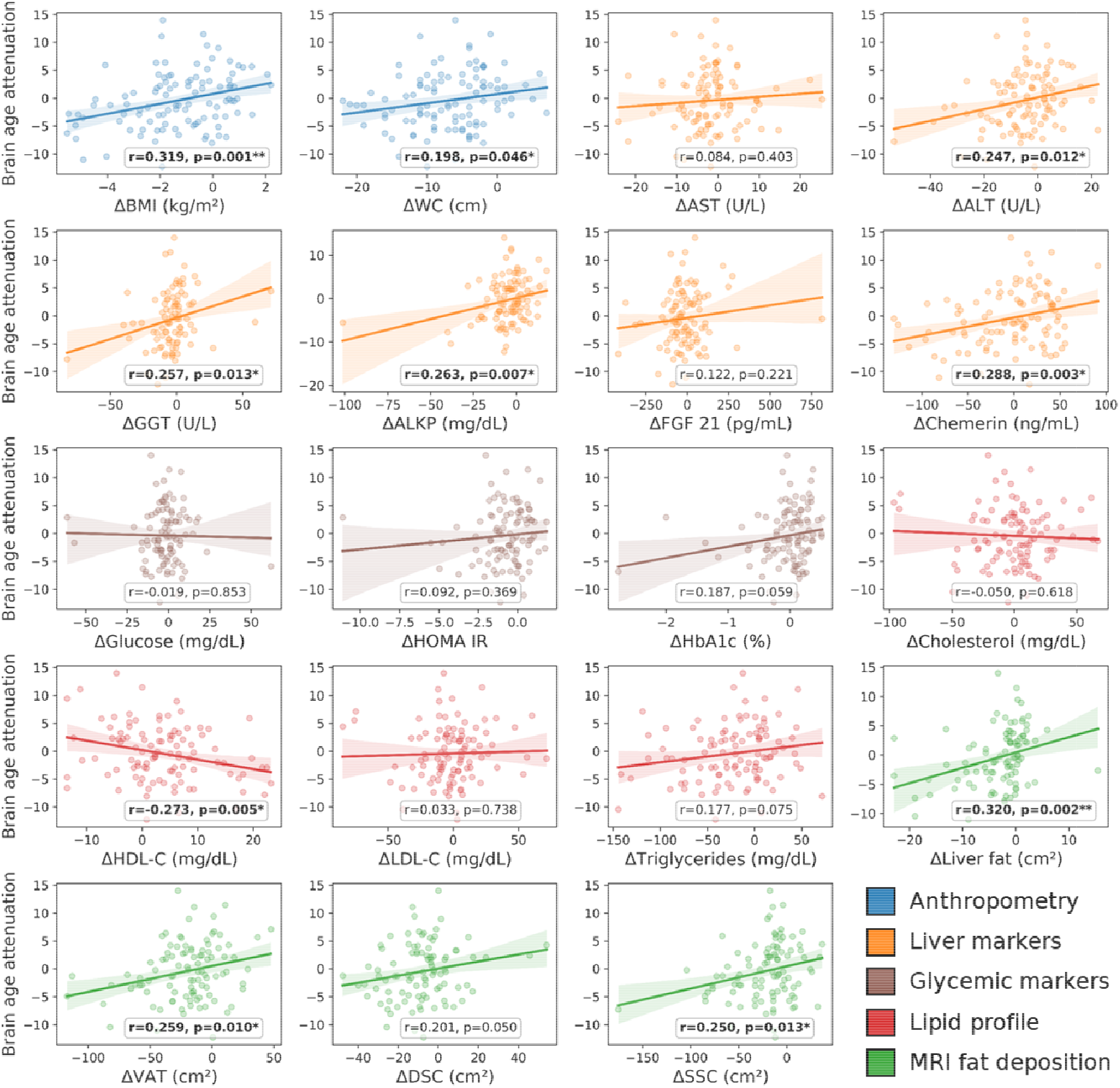
Brain age attenuation association with clinical measurements. The scatter plots depict the data points and regression line between brain age attenuation (y-axis) and each clinical measurement (x-axis). Clinical measurements include anthropometry (blue), liver markers (orange), glycemic markers (brown), lipid profile (red), and fat deposition measured using MRI (green). The shaded area around the regression line represents a 95% confidence interval estimated using bootstrapping. Pearson’s correlation and the corresponding p-value are shown at the bottom of each plot. Significant associations following FDR correction are marked in bold (*=p<0.05, **=p<0.01).

### 2.4 The relation between brain age attenuation and clinical measurements

To examine the clinical outcomes associated with attenuated brain aging, we further tested the correlation of brain age attenuation with liver, glycemic, lipids, and MRI-assessed fat deposition biomarkers (Fig. 4). Except of deep subcutaneous changes, all fat deposition measurements, superficial subcutaneous, visceral, and liver fat changes (e.g loss) were significantly and directly associated with brain age attenuation (p<0.05, FDR corrected), i.e., the more the individual succeeded in diet-induced fat depots loss, the more brain age attenuation has been achieved. We then tested the association between brain age attenuation and liver and glycemic biomarkers. Out of all examined liver biomarkers, a decrease in alanine transaminase (ALT), Gamma-glutamyl Transferase (GGT), alkaline phosphatase, and serum chemerin were significantly associated with attenuation in brain age (p<0.05 for all, FDR corrected). Of all examined lipid profile markers, only an increase in ΔHDL-C was significantly correlated with brain age attenuation (r=-.273, p=0.005). Finally, a decrease in HOC was significantly correlated with brain age attenuation (r=-.296, p=0.003). All results were reproduced after controlling for baseline age and predicted age at T0. However, after further correction for changes in BMI, only Δ alkaline phosphatase, Δ chemerin and Δ HOC were associated with brain age attenuation (all p’s<0.018), with no significant associations with all other biomarkers (p>0.05, for all; SI Table 2).

### 2.5 Relation between brain age attenuation and food consumption

We examined whether food consumption, as reported using a Food Frequency Questionnaire (FFQ), could be associated with functional brain aging attenuation. Associations were tested using Kendall’s rank correlation. We began with categories that could negatively affect brain aging attenuation. In line with our hypothesis, we found that decreased consumption of processed food (t=3.131, p=.002) and sweets and beverages (τ=-0.231, p=.002) was associated with more attenuation in brain age. An increase in green tea and walnut consumption, for which we hypothesized an attenuation effect on brain aging due to their high polyphenol content, did not result in a significant correlation (all τ’s <0.081, p’s>.121).

## 3. Discussion

Considerable evidence implies that excessive weight accelerates normal aging^11,12^, a process that is also manifested in brain aging^38^. In the current study, we examined for the first time, as far as we can say, whether a lifestyle dietary intervention may attenuate the effect of obesity on the brain aging trajectory. We hypothesized that reducing anthropometric measurements following a lifestyle intervention would be associated with attenuated brain aging. We first demonstrated, across two separate cohorts, that age could be estimated from RSFC, as done in previous work^39^. We then applied the fitted age prediction model to the participants of the DIRECT-PLUS. We found that 1% of body weight loss results in an 8.9 months attenuation of brain age (SI Fig. 2). Attenuated brain aging was further correlated with a decrease in WC, MRI assessed fat deposition, liver biomarkers, and HDL-C. Finally, reduced reported consumption of processed food, sweets and beverages were also related to attenuated brain aging.

Accumulated evidence points to the potential of lifestyle intervention to reverse the negative impact of excess weight on brain structure, function, and cognition. Cross-sectional and longitudinal studies found that reported adherence to a Mediterranean diet was linked to increased gray matter volume in multiple regions^40^, including the hippocampus^41^. Adherence to healthy dietary patterns was also associated with reduced cognitive decline^42^. Importantly, randomized clinical trials can support a causal relationship between lifestyle intervention and the brain aging process. Such studies from our group^32^ and others^43,44^ revealed that subjects enrolled in a PA + dietary intervention exhibit lower hippocampal atrophy and smaller ventricles. A similar beneficial effect on cognitive functioning in middle age was also found^45^, along with functional connectivity alteration in the default mode and executive control networks^46,47^. To date, a single study in rats has tested the effect of a dietary intervention using the brain age framework and found a reduction in brain aging rate^48^. Hence, the current work provides the first evidence that such a beneficial effect on brain age can also be found in humans.

The brain age framework reduces the multifaceted aging process captured in a given imaging modality to a single scalar. This scalar, the predicted brain age, is well-defined in the sense that it minimizes the prediction error within the training data set. Moreover, the clinical relevance of functional brain age is shown, for example, in predicting Alzheimer’s onset^49^ and symptoms severity in depression^50^. This reductionist approach raises several challenges. The first is the ability to interpret the features used by the machine learning model^51^ (see SI 1). A second challenge is understanding the physiological factors that may affect its predictions^52^, which we address in the current work. Here we report how a set of clinical measures are associated with changes in brain aging. Importantly, lifestyle and other interventions can affect these measures to attenuate the brain aging process. We suggest that such mapping of changes in clinical outcomes to months or years of attenuated brain aging has important scientific, clinical, and even educational value.

We found that clinical outcomes that include anthropometric, liver, and lipid biomarkers were associated with attenuated brain age. Specifically, two main factors were linked to changes in brain age, changes in anthropometry measures, and liver status. The first factor included BMI, weight, WC, and superficial subcutaneous and visceral fat. The second factor included liver fat, ALT, GGT, alkaline phosphatase, and serum chemerin. Alkaline phosphatase and chemerin were also associated with changes in brain age after controlling for changes in BMI. The negative impact of elevated liver enzymes and liver fat on brain health is seen, for example, in the case of AD^53–55^. This link is thought to be mediated by oxidative stress, vascular damage, and inflammation^56^. Chemerin, produced in the liver, is an adipokine linked to energy homeostasis, adipogenesis, and excessive weight^56^. Chemerin is correlated with age^57^and BMI^58^ and was found to be reduced following lifestyle intervention^59,60^. The relation between serum chemerin and brain aging is still unclear, but possible linking mechanisms are hypertension^61^ and inflammation^58^. Besides these two factors, HDL-C was the only variable whose increase was correlated to brain aging attenuation. This is in line with evidence of the protective role of HDL-C in cognitive decline and dementia^62^. Finally, of all the reported food consumption items, only reduced consumption of processed food, sweets and beverages were linked to attenuated brain aging. Although these results are based on self-reports, they may be helpful for developing neuroprotective dietary guidelines^63^.

It is important to consider several limitations and strengths of the current study. The first limitation was gender imbalance (F: 93, M: 9; F:8.8%, M: 91.2%), which reflected the workplace profile from which participants were recruited^64–67^. This distribution misrepresents the proportion of obese women within the general population (F: 51%, M: 49%; CDC, 2018). Hence these results should be further corroborated in a gender-balanced sample. Additionally, participants were recruited based on excess adiposity or dyslipidemia, therefore, they represented a restricted range of the normal population. This design allows to maximize the intervention effects but restricts our ability to detect correlation at baseline. The strengths of the study included the wealth of health biomarkers that included anthropometric, blood, and imaging measures. The relatively large sample for similar intervention trials, the tight on-site monitoring over the dietary compliance, and the long intervention duration. Finally, the use of three distinct datasets for training and validation, testing, and inference supports the generalization of our model.

To conclude, in the current work, we examined how changes in multiple health factors, including anthropometric measurements, blood biomarkers, and fat deposition, can account for brain aging attenuation. We reveal that the two factors with the strongest association with brain aging were changes in anthropometry measures and liver biomarkers. These findings complement the growing interest in bodily aging indicated, for example, by DNA methylation^68^ as health biomarkers and interventions that may affect them. These exciting results may advance our knowledge of factors related to healthy brain aging and guide future neuroprotective interventions.

## 4. Methods

### 4.1 Dataset used for training and validating the brain age model

Training, validation, and testing of the brain age model were conducted on data from two cohorts that included functional and structural brain magnetic resonance imaging (MRI). The training was conducted on the enhanced Nathan Kline Institute-Rockland Sample (NKI^34^) and testing on the Cambridge Centre for Ageing and Neuroscience dataset (Cam-CAN^35,36^). The NKI dataset is composed of 291 subjects (226 females, 65 males) recruited from Rockland County, USA. All participants provided informed consent and the study was approved by the Institutional Review Board at the Nathan Kline Institute (#226781 and #239708) and Montclair State University (#000983 A and #000983B). The Cam-CAN dataset includes 358 (193 females, 165 males) subjects roughly uniformly distributed from Cambridge City, UK. All participants provided informed consent, and the study was approved by the local ethics committee, Cambridgeshire 2 Research Ethics Committee (reference: 10/H0308/50). In both datasets, we included only subjects within the DIRECT-PLUS age range (34-82 years). Exclusion criteria included unsuccessful completion of the preprocessing and quality control stages (see 4.5).

### 4.2 Study design

This work was based on a sub-study of the DIREC-PLUS trial (clinicaltrials.gov ID: NCT03020186). The primary aims of the DIRECT-PLUS trial were 18-month changes in visceral abdominal tissue, intrahepatic fat, and adiposity across intervention groups. The results for the primary outcomes were presented in separate publications^33^. The DIRECT-PLUS was launched in May 2017 and conducted in an isolated workplace in Israel (Nuclear Research Center Negev, Dimona, Israel). Most clinical and medical measurements, including anthropometric measurements, blood drawing, and lifestyle intervention sessions, were performed on-site. Among 378 volunteers, 294 met age (30+ years of age) and abdominal obesity inclusion criteria [waist circumference (WC): men>102 cm, women>88 cm] or dyslipidemia [TG>150 mg/dL and high-density-lipoprotein-cholesterol (HDL-c) ≤40 mg/dL for men, ≤50 mg/dL for women]. Exclusion criteria were inability to perform physical activity; serum creatinine ≥2 mg/dL; serum alanine aminotransferase or aspartate aminotransferase more than three times above the upper limit of normal; a major illness that might require hospitalization; pregnancy or lactation; active cancer, or chemotherapy treatment in the last three years; warfarin treatment; pacemaker or platinum implantation; and participation in a different trial. Among 294 eligible participants, 132 participants were randomly assigned to participate in the fMRI sub-study. The Soroka Medical Center Medical Ethics Board and Institutional Review Board provided ethics approval. All participants provided written consent and received no financial compensation.

### 4.3 Randomization and intervention

All participants completed the baseline measurements and were randomized, using a computer-basd program, in a 1:1:1 ratio, stratified by sex and work status (to ensure equal workplace-related lifestyle features between groups), into one of the three intervention groups: healthy dietary guidelines (HDG) as an active control group, Mediterranean diet (MED), green-MED, all combined with physical activity (PA). Interventions lasted for 18 months and were contemporaneous, and participants were not blind to group assignment (open-label protocol). Each participant received complete dietary guidance (based on the specific intervention group) and a free and fully available clinical dietitians consult. Furthermore, all participants received free gym membership, including educational sessions encouraging moderate-intensity PA. A detailed description of the intervention outline is available in **SI Table 3**.

### 4.4 MRI acquisition

MRI scans were conducted at the Soroka University Medical Center (SUMC), Beer Sheva. Participants were scanned in a 3T Philips Ingenia scanner (Amsterdam, The Netherlands) equipped with a standard head coil. Subjects were instructed to refrain from food and non-water beverages two hours before the MRI sessions. Each of the two sessions at T0 and T18 included 2 RS-fMRI runs of 7 minutes each and a 3D T1-weighted anatomical scan to allow registration of the functional data. Before each RS session, participants were instructed to remain awake with their eyes open and lie still. fMRI BOLD contrast was acquired using the gradient-echo echo-planner imaging sequence with parallel acquisition (SENSE: factor 2.2). Scanning parameters were as follows: whole-brain coverage 41 slices (3 × 3 × 3 mm3), transverse orientation, 3 mm thickness, no gap, TR = 2200 ms, TE = 30 ms, flip angle=90°, FOV = 200 × 222 (RL x AP) and matrix size 68 × 71 (RL x AP). High-resolution anatomical volumes were acquired with a T1-weighted 3D pulse sequence (1 × 1 × 1 mm3, 150 slices). A detailed description of liver and abdominal fat MRI is available in SI section 10.2.

### 4.5 MRI preprocessing

The preprocessing pipelines used in this work were extensively described in a previous publication^69^. T1w scans were preprocessed through FreeSurfer’s^70^ (version 6.0) recon-all processing. FreeSurfer’s cortical segmentation and spherical warp were used to transfer the Schaefer 100-node cortical parcellation^37^ to each subject’s volumetric anatomical space. Functional images of the NKI dataset were preprocessed with fMRIPrep (version 1.1.8;^71^ and images of the DIRECT-PLUS and Cam-CAN datasets were preprocessed with the Configurable Pipeline for the Analysis of Connectomes (C-PAC^72^ version 1.6.2). Briefly, both pipelines included the following steps: slice-timing correction, motion correction, skull stripping, estimation of motion parameters, and other nuisance signal time series. For the NKI dataset, functional scans were bandpass filtered (0.008 – 0.08Hz) and confound regressed in a manner orthogonal to the temporal filters. Confounds included six motion estimates, the mean time series derived in CSF, WM, and whole-brain masks, the derivatives of these nine regressors, and the squares of these 18 terms. Spike regressors were added for each frame with framewise displacement above 0.5mm. Data were linearly detrended and standardized. Nuisance regression in the DIRECT-PLUS and Cam-CAN fMRI dataset included the first five principal components of the signal from white matter and CSF^73^, six motion parameters, and linear and quadratic trends, global signal regression, followed by temporal filtering between 0.1 and 0.01 Hz and. Finally, a scrubbing threshold of 0.5mm frame-wise displacement was applied^74^ (removal of 1 TR before and 2 TR after excessive movement). The time series of the two functional scans in the DIRECT-PLUS were concatenated to a single T0 and T18 scans. The exclusion criterion for excessive movements was determined a priori to less than 70% (9 min and 48 sec) of the resting-state session after the scrubbing procedure (23% omitted; 102 subjects left). In all datasets, functional connectivity was defined as the Pearson correlation among pairs of ROIs’ time series followed by Fisher’s r-to-z transformation.

### 4.6 Clinical measurement and blood biomarkers

All parameters were measured at baseline and after 18 months of intervention. Waist circumference was measured to the nearest millimeter halfway between the last rib and the iliac crest using an anthropometric measuring tape. Blood and urine samples were collected at 8:00 AM after a 12-hour fast. Blood samples were centrifuged and stored at -80°C. Hippocampal occupancy score (HOC) was calculated as the hippocampal volume divided by [hippocampal volume + inferior lateral ventricle volume] in each hemisphere, then averaged across hemispheres ^32,75^.

### 4.7 Nutritional assessment

Assessment of nutritional intake and lifestyle habits was self-reported online using validated food frequency questionnaires^76^ including green tea, walnuts, and *Wolffia globose* intake evaluation. The questionnaires were administered at baseline, after six months, and at the end of the trial. The closed workplace enabled monitoring of the freely provided lunch and the intense dietary and PA sessions, which were provided simultaneously to all three groups.

### 4.8 Brain age estimation

Subjects’ chronological age was predicted from the lower triangle of the functional connectivity matrix depicting all unique edges (4950 edges). We used a support vector regression model^77^ implemented using Scikit-learn^78^ with a linear kernel and all the default parameters. Model accuracy was quantified as the Pearson correlation between the observed and predicted age. We additionally report the mean absolute error (MAE) in years, along with a p-value based on a non-parametric permutation test created by shuffling the data labels 1,000 times^79^.

### 4.9 Statistical analysis

The primary outcome of the current work was brain age attenuation quantified as the difference between the expected and observed brain age at T18. The expected brain age at T18 was calculated by first producing brain age prediction for all participants at T0. Then, a linear regression was used to estimate brain age from the chronological age at T0. The fitted regression formula, representing the expected aging trajectory in the absence of intervention, was used to estimate the expected brain age at T18 given each participant’s T0 brain age and the time passed between the T0 and T18 MRI scans. The observed brain age was produced by applying the brain age model to the T18 scans. Association between brain age attenuation and change in clinical measures following the intervention were reported using Pearson’s correlation. Correction for multiple comparisons was conducted within each biomarker category using the Benjamini– Hochberg false discovery rate (FDR^80^) with an alpha of 0.05. Associations to food consumption reports were reported using Kendall’s tau correlation for ordinal data. Processed food at T18 had only two levels, “same consumption” and “less consumption”, thus relation to brain age attenuation was tested with independent t-test. Change in clinical measurements were computed as a delta (Δ), the value at T0 minus the value at T18. We quantified change in reported food consumption as the change between the T0 and T18 questionnaires for food groups (i.e., processed food, sweets, and beverages) and as total consumption for polyphenols-provided foods (i.e., Mankai, green tea, walnuts). To control for the possible effect of age or gender, we used partial regression by regressing out the linear effect of age and gender from both brain age attenuation and the clinical measures. This was done by predicting each clinical measure, with the covariate as a predictor, keeping only the residual.

## Data Availability

The data are not publicly available due to ethical and privacy restrictions.

## 5. Author contributions

G.L.: Conceptualization, Methodology, Investigation, Formal analysis, Software, Visualization, Writing - original draft, Writing - review & editing. A.K.: Methodology, Investigation, Formal analysis, Software, Project administration, Writing - original draft, Writing - review & editing. A.Y.M.: Investigation, Project administration. E.R.: Investigation, Project administration. G.T.: Investigation, Project administration. H.Z.: Investigation, Project administration. I.S.: Resources, Conceptualization, Methodology, Writing - review & editing. G.A.: Conceptualization, Methodology, Writing - original draft, Writing - review & editing, Supervision. I.S.: Resources, Conceptualization, Methodology, Supervision, Funding acquisition, Writing - review & editing; PI of the DIRECT-PLUS. The ordering among co–first authors was determined by their relative contributions to the study.

## 6. Acknowledgments

This work was supported by grants from: the German Research Foundation (DFG), German Research Foundation - project number 209933838 - SFB 1052; B11), Israel Ministry of Health grant 87472511 (to I Shai); Israel Ministry of Science and Technology grant 3-13604 (to I Shai); and the California Walnuts Commission (to I Shai).

## 10. Supplemental material

### 10.1 Brain age prediction accuracy of individual nodes

To examine the contribution of different nodes for age prediction, we reiterated the model training procedure for each of the nodes separately. We extracted the row of each node in the RSFC, matrix which represents all the nodes’ connections with the rest of the brain, or its ‘connectivity fingerprint’^81^. As in the original model, we used a linear support vector regression fitted on the NKI data set, then tested it on the Cam-CAN data set. We report the prediction accuracy of each node on the test set as the Pearson correlation between the predicted brain age and the chronological age. We plot the results on the brain surface and using a box plot where nodes are arranged according to their resting-state network affiliations (SI fig. 1).

**SI Fig.1.**
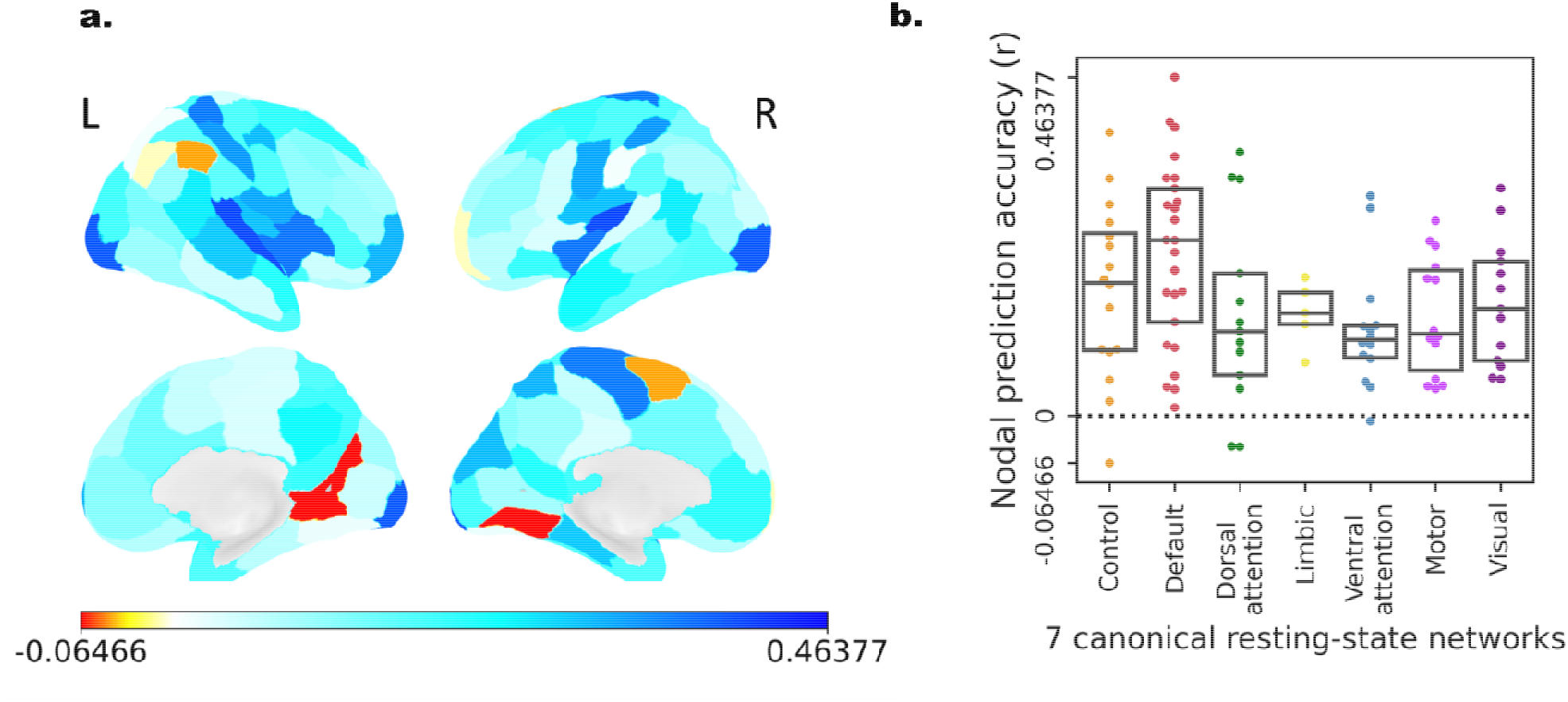
Brain age prediction accuracy of individual nodes. Prediction accuracy was quantified as the correlation between chronological age and predicted age. (a) Prediction accuracy was depicted on the brain surface on a lateral (top) and medial (bottom) view. (b) Box plot showing the prediction accuracy (y-axis) in each of the seven canonical resting-state networks ^82^. Each dot represents a single node.

**SI Table 1.**
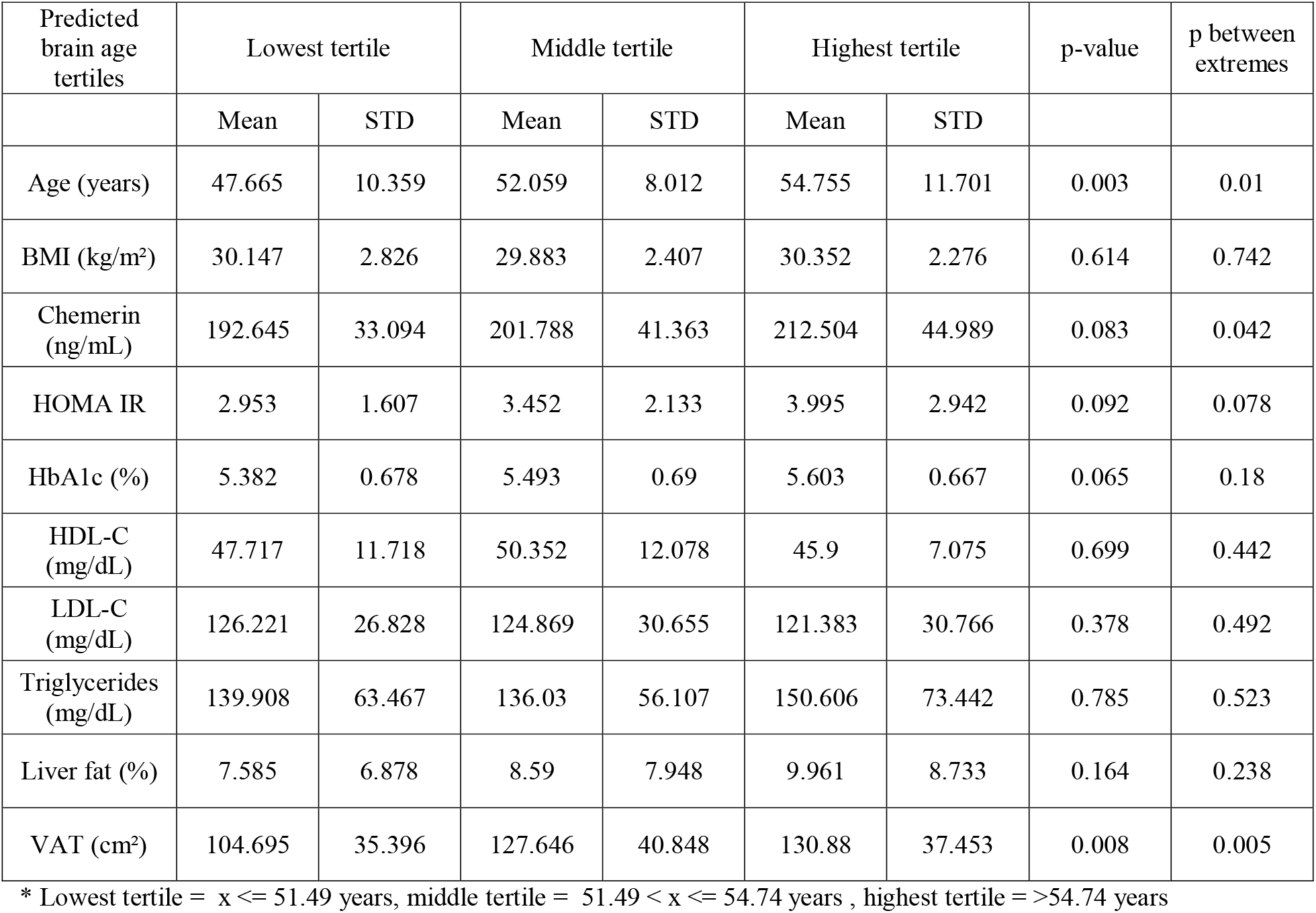
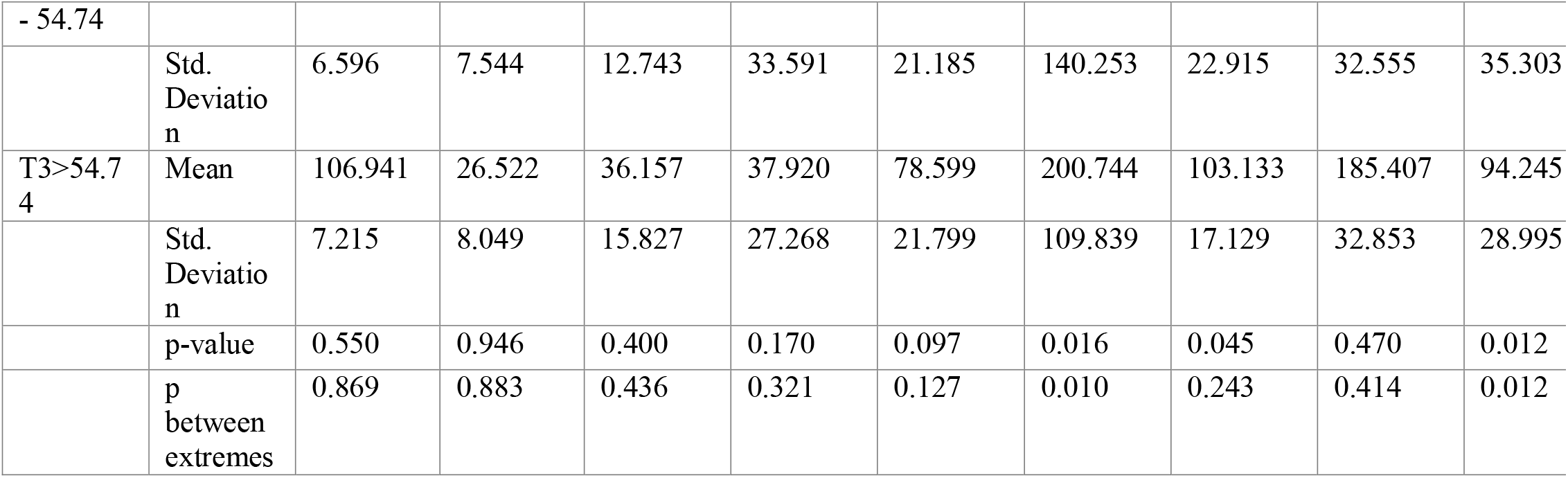
Baseline characteristics according to the baseline predicted brain age tertiles

**SI Fig. 2.**
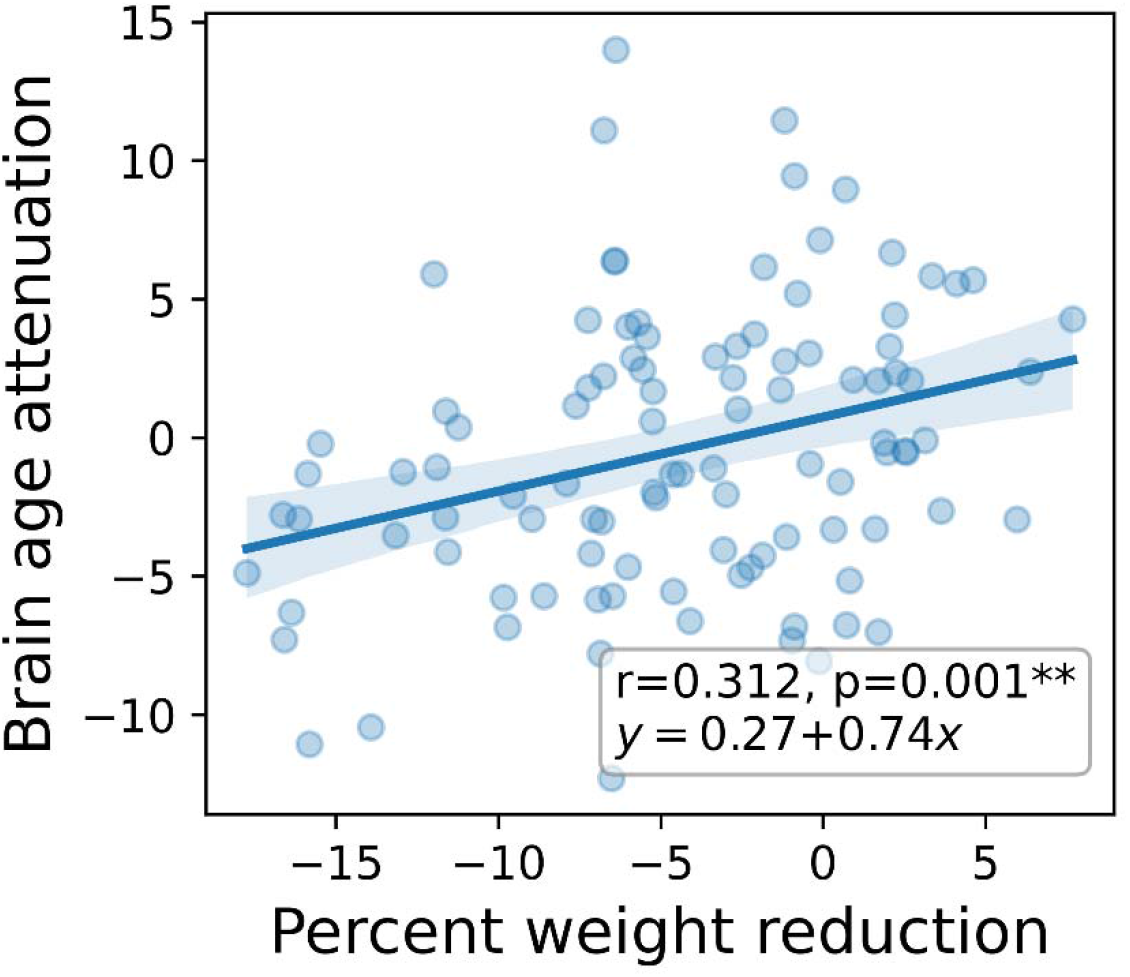
Brain age attenuation compared to percent weight reduction from baseline. A scatter plot depicting the linear relationship between percent weight reduction from baseline (x-axis) and years of brain age attenuation (y-axis). Also shown on the graph are the correlation coefficient and the parameters of the linear relation between the two variables. One percent of body weight loss corresponded to an. months attenuation of brain age.

**SI Table 2.** Correlation and partial correlation of brain age attenuation and biomarkers.

|  | r | p-value | r (age-corrected) | p-value (age-corrected) | r (baseline brain age - corrected) | p-value (baseline brain age-corrected) | r (ΔBMI-corrected) | p-value (ΔBMI-corrected) |
| --- | --- | --- | --- | --- | --- | --- | --- | --- |
| ΔBMI (kg/m<sup>2</sup>) | **0.319** | **0.001** | **0.319** | **0.001** | **0.266** | **0.007** | -0.041 | 0.682 |
| ΔWC (cm) | **0.198** | **0.046** | **0.198** | **0.046** | 0.171 | 0.085 | -0.042 | 0.674 |
| ΔAST (U/L) | 0.084 | 0.403 | 0.092 | 0.359 | 0.053 | 0.599 | 0.052 | 0.603 |
| ΔALT (U/L) | **0.247** | **0.012** | **0.268** | **0.007** | **0.197** | **0.047** | 0.135 | 0.176 |
| ΔGGT (U/L) | **0.257** | **0.013** | **0.264** | **0.011** | **0.237** | **0.022** | 0.157 | 0.133 |
| ΔALKP (mg/dL) | **0.263** | **0.007** | **0.281** | **0.004** | **0.230** | **0.020** | **0.233** | **0.018** |
| ΔFGF 21 (pg/mL) | 0.122 | 0.221 | 0.123 | 0.217 | 0.094 | 0.346 | 0.044 | 0.659 |
| ΔChemerin (ng/mL) | **0.288** | **0.003** | **0.290** | **0.003** | **0.219** | **0.027** | **0.245** | **0.013** |
| ΔGlucose (mg/dL) | -0.019 | 0.853 | -0.005 | 0.958 | -0.101 | 0.323 | -0.088 | 0.389 |
| ΔHOMA IR | 0.092 | 0.369 | 0.107 | 0.293 | -0.018 | 0.862 | -0.027 | 0.789 |
| ΔHbA1c (%) | 0.187 | 0.059 | **0.198** | **0.046** | 0.164 | 0.099 | 0.090 | 0.371 |
| ΔCholesterol (mg/dL) | -0.050 | 0.618 | -0.048 | 0.629 | -0.008 | 0.934 | -0.020 | 0.840 |
| ΔHDL-C (mg/dL) | **-0.273** | **0.005** | **-0.272** | **0.006** | **-0.228** | **0.021** | -0.144 | 0.149 |
| ΔLDL-C (mg/dL) | 0.033 | 0.738 | 0.035 | 0.730 | 0.069 | 0.493 | 0.060 | 0.551 |
| ΔTriglycerides (mg/dL) | 0.177 | 0.075 | 0.180 | 0.071 | 0.166 | 0.095 | 0.082 | 0.412 |
| ΔLiver fat (cm²) | **0.259** | **0.010** | **0.264** | **0.008** | **0.225** | **0.025** | 0.045 | 0.659 |
| ΔVAT (cm²) | **0.320** | **0.002** | **0.334** | **0.001** | **0.297** | **0.004** | 0.178 | 0.085 |
| ΔDSC (cm²) | 0.201 | 0.050 | **0.202** | **0.049** | **0.215** | **0.037** | -0.024 | 0.818 |
| ΔSSC (cm²) | **0.250** | **0.013** | **0.246** | **0.014** | **0.232** | **0.022** | 0.007 | 0.947 |
| ΔHOC | **-0.296** | **0.003** | **-0.294** | **0.003** | **-0.346** | **<0.001** | **-0.269** | **0.007** |

\* All delta values were calculated as the value at T0 minus the value at T18

**SI Table 3.**
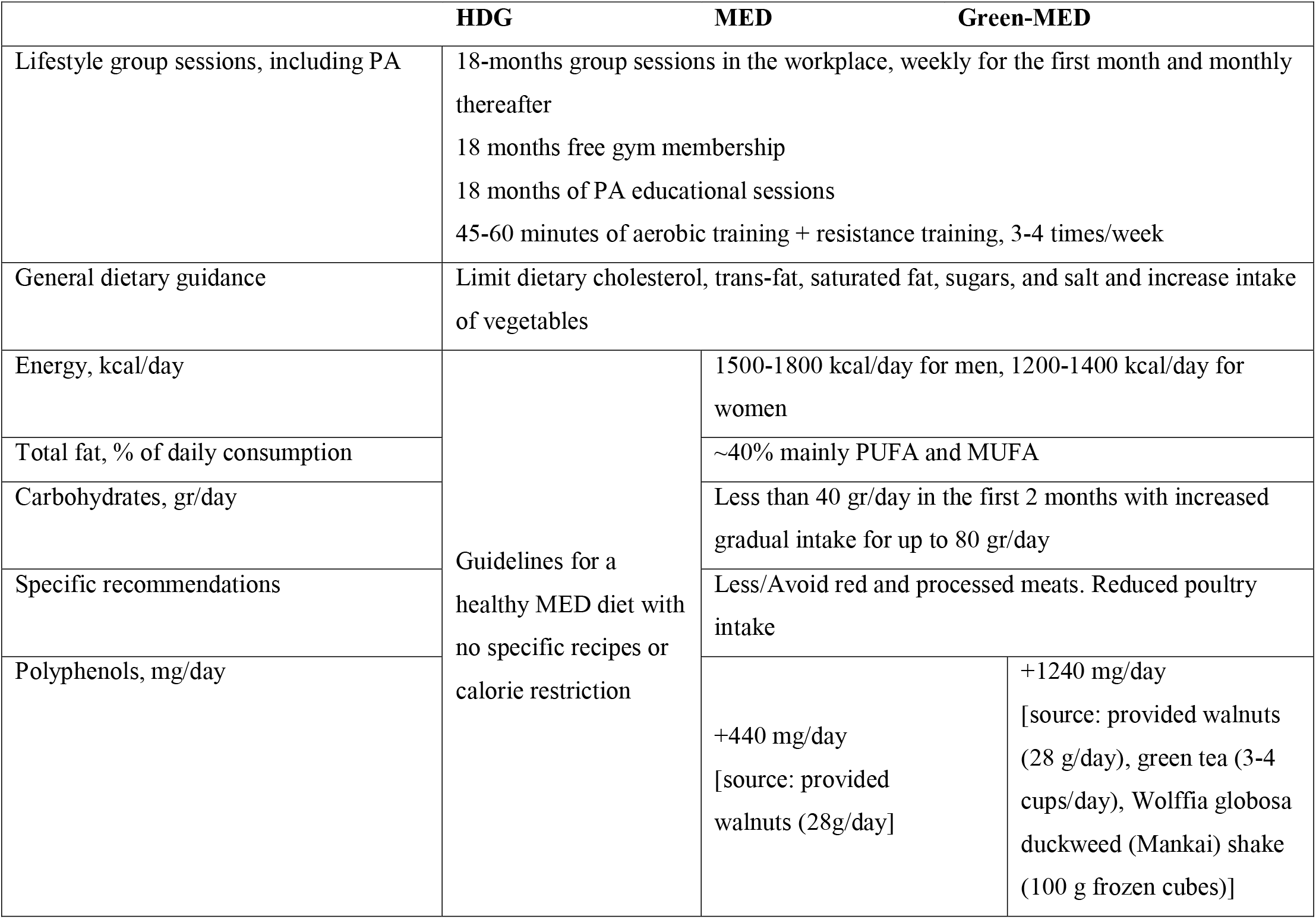
Intervention outline by group.

|  | HDG | MED | Green-MED |
| --- | --- | --- | --- |
| Lifestyle group sessions, including PA | 18-months group sessions in the workplace, weekly for the first month and monthly thereafter<br>18 months free gym membership<br>18 months of PA educational sessions<br>45-60 minutes of aerobic training + resistance training, 3-4 times/week |  |  |
| General dietary guidance | Limit dietary cholesterol, trans-fat, saturated fat, sugars, and salt and increase intake of vegetables |  |  |
| Energy, kcal/day | Guidelines for a healthy MED diet with no specific recipes or calorie restriction | 1500-1800 kcal/day for men, 1200-1400 kcal/day for women |  |
| Total fat, % of daily consumption |  | ~40% mainly PUFA and MUFA |  |
| Carbohydrates, gr/day |  | Less than 40 gr/day in the first 2 months with increased gradual intake for up to 80 gr/day |  |
| Specific recommendations |  | Less/Avoid red and processed meats. Reduced poultry intake |  |
| Polyphenols, mg/day |  | +440 mg/day<br>[source: provided walnuts (28g/day)] | +1240 mg/day<br>[source: provided walnuts (28 g/day), green tea (3-4 cups/day), Wolffia globosa duckweed (Mankai) shake (100 g frozen cubes)] |

### 10.2 Liver and visceral fat imaging protocols

To quantify and follow IHF% changes, we used H-MRS, a reliable tool for liver fat quantification (https://pubmed.ncbi.nlm.nih.gov/25903702/). Localized, single-voxel proton spectra were acquired using a 3.0T magnetic resonance scanner (Philips Ingenia, Best, The Netherlands). The measurements were taken from the right frontal lobe of the liver, with a location determined individually for each subject using a surface, receive-only phased-array coil. Spectra with and without water suppression were acquired using the single-voxel stimulated echo acquisition mode (STEAM) with the following parameters: TR=4000msec, TE=9.0msec and TM=16.0msec. The receiver bandwidth was 2000Hz and the number of data points was 1024. Second-order shimming was used. Four averages were taken in a single breath hold for an acquisition time of 16 sec. The voxel size varied somewhat according to anatomy, but was approximately 50(AP) × 45(RL) × 54(FH) mm. Water suppression was achieved using the MOIST (Multiple Optimizations Insensitive Suppression Train) sequence consisting of four phase-modulated T1 and B1 insensitive pulses with a 50Hz window. Data were analyzed using Mnova software (Mestrelab Research, Santiago de Compostela, Spain) by an experienced physicist blinded to the intervention groups, who also performed visual quality control of fitted spectra. The total image hepatic fat fraction was determined as the ratio between the sum of the area under all fat divided by the sum of area under all fat and water peaks (Cite: https://pubmed.ncbi.nlm.nih.gov/19834463/). Inter-class reliability was tested between two different technicians and resulted in an average measure of r=0.99 (p<0.001). Intra-class reliability was tested among all baseline scans and resulted in an average measure of r=0.96 (p<0.001). Liver fat color images were produced using PRIDE software (by Philips).

Abdominal fat depots were assessed at baseline and 18-months thereafter using 3-Tesla MRI scans (Ingenia 3.0T, Philips Healthcare, Best, the Netherlands). The scanner utilized a 3D modified DIXON (mDIXON) imaging technique without gaps (2 mm thickness and 2 mm of spacing), fast-low-angle shot (FLASH) sequence with a multi-echo two excitation pulse sequence for phase-sensitive encoding of fat and water signals (TR, 3.6ms; TE1,1.19ms; TE2, 2.3ms; FOV 520×440×80mm; 2×1.4×1mm voxel size). Four images of phantoms were generated: in-phase, out-phase, fat, and water phase ^83^. Participants were instructed to hold their breath to avoid motion artifacts when their abdomen was scanned. We quantified abdominal fat using MATLAB-based semiautomatic software and blinded to intervention group. A continuous line over the fascia superficialis was drawn to differentiate deep-SAT from superficial-SAT and calculated mean VAT, deep-SAT, and superficial-SAT along two axial slices: L5-S1 and L4-L5. We quantified fat mass regions as area and relative proportion of each fat subtype (percentage).

